# Spinal Sarcopenia in Patients with Spinal Osteoarthritis Across Different Age Groups: Clinical and MRI Characteristics

**DOI:** 10.1101/2025.02.11.25321878

**Authors:** N.G. Pravdyuk, A.V. Novikova, N.A. Shostak, A.A. Klimenko, E.S. Pershina, A.A. Muradyants, A.A. Buianova

## Abstract

**Introduction:** Sarcopenia of the spinal stabilizing muscles is one of the components of age-related sarcopenia and often associated with chronic back pain (CBP), including spinal osteoarthritis (OA). The age dynamics of paraspinal muscle mass in spinal OA is not sufficiently studied to date, although it is key in developing rehabilitation and prevention strategies.

**The aim of study:** is to identify spinal sarcopenia in individuals with spinal OA by assessing paraspinal muscles mass and analyzing its relationship to degenerative spinal changes in different age groups using magnetic resonance imaging (MRI).

**Materials and methods:** The study included 90 patients with spinal OA classified as stages 4 and 5 of degenerative disc disease according to the Pfirrmann grading system. Participants were evenly distributed across three age groups: young adults (Me: 36.00 [30.00-42.00]), middle age (Me: 50.00 [46.50-56.25]), and elderly (Me: 66.00 [62.50-71.00]). All participants underwent lumbar spine MRI in two sections – sagittal and axial – in T1- and T2-weighted images.

Clinical assessments included evaluating the functional status of the lumbar spine using the Backache Index (BAI). Paraspinal muscle mass analysis was performed on axial sections at the level of the third lumbar vertebra (L3) for the following muscles: musculus psoas major, musculus erector spinae, and musculus quadratus lumborum. To comparatively assess spinal muscle mass, the musculovertebral index (MVI) was calculated by dividing the total cross-sectional area of the three pair muscles (Sm, cm²) by the cross-sectional area of the third lumbar vertebra (Sv, cm²) at the upper endplate level: MVI = (Sm dextra + Sm sinistra) / Sv.

To identify the asymmetries of the paraspinal muscles, the ratio of the cross-sectional areas of the three muscles on the left and right sides was calculated. Additionally, the transverse torso thickness at L3 level was measured on sagittal sections.

**Results:** The mean MVI values demonstrated that paraspinal muscle mass was significantly lower in elderly (p = 0.0001) and middle-aged (p = 0.025) groups compared to the young group, which indicates its decrease with age in patients.

Statistically significant increase in the lumbar vertebra area with age (p=0.0003 young vs elderly) is explained by the spondylosis development and age-related marginal osteophyte growth as manifestations of spinal OA.

The reduction in muscle cross-sectional area was evident for three right paraspinal muscle groups with age (p = 0.0252). Asymmetry for each pair of muscles was observed across all age groups, the right side always being larger than the left, particularly in the quadratus lumborum, where it was most pronounced in middle-aged (p = 2.762 × 10^−6^) and young individuals (p = 0.0009).

The elderly persons demonstrated a tendency toward higher body mass index, a higher percentage of obese individuals, and increased torso thickness compared to the middle-aged and young groups (p = 0.067), suggesting a possible decline in abdominal muscle tone with age alongside body weight gain.

**Conclusions:** MRI findings reveal an age-related decline in paraspinal muscle mass among patients with CBP, indicating the development of spinal sarcopenia, which likely contributes to the clinical course of spinal OA in the elderly. All three groups of muscles show asymmetry in all age groups, with the right side always being larger than the left.

Torso thickness and body weight increase with age in patients with spinal OA potentially exacerbate the disease’s clinical progression. These findings highlight the need to include exercises targeting paraspinal and abdominal muscles in rehabilitation programs for patients with spinal OA in all age groups. Preventing spinal spinal sarcopenia by maintaining axial muscle stability is essential for preserving the spinal muscle framework and is an integral component of managing patients with CBP.

## Introduction

Spinal osteoarthritis (OA) is one of the leading age-associated conditions resulting from degenerative changes in the musculoskeletal system. The prevalence of chronic back pain (CBP) associated with OA has risen over recent decades across both sexes and tends to peak in the eighth decade of life [1]. Pain in spinal OA, akin to OA of peripheral joints, involves all structures of the spinal functional unit, including the intervertebral disc, the three-joint complex of the vertebral motion segment, and the associated musculoligamentous apparatus. Among all components of the functional spinal unit (FSU), the muscular framework remains the least studied in the context of spinal degeneration. It is well-established that trunk muscles – comprising the back, abdominal, pelvic diaphragm, and lower limb muscles – constitute the core stabilizing the spine and governing its movements through coordinated activity [2,3]. The paraspinal muscles, which are in direct anatomical connection with the vertebral motion segments, play a critical role in regulating spinal motion and ensuring stability [4]. This interdependence is especially significant in CBP, as paraspinal muscles help prevent excessive vertebral displacement, pathological curvatures, and critical intervertebral disc deformation [5].

CBP has a dual impact: it triggers compensatory spasms aimed at stabilizing the affected spinal segments while simultaneously reducing paraspinal muscle activity due to restricted lumbar mobility, ultimately leading to the loss of active myofibrils. This phenomenon of muscle mass and strength reduction, well-documented in elderly populations, is termed ‘sarcopenia’. Sarcopenia refers to a progressive loss of skeletal muscle mass and strength, often accompanied by diminished functional activity [6]. Spinal sarcopenia may manifest as a primary age-related process or as a secondary condition associated with localized spinal pathology, such as intervertebral disc degeneration. This is analogous to localized muscle mass loss observed in immobilized limbs following prolonged disuse. Recurrent CBP is marked by episodic spinal immobilization and compensatory paraspinal muscle spasms aimed at stabilizing affected segments [7]. Furthermore, in the advanced stages of spinal OA, significant damage to the intervertebral disc with its subtotal dehydration and a height reduction of more than 50% (stage 4-5 according to Pfirrmann) creates additional reasons for immobilization of the intervertebral joints and the entire lumbar spine, which contributes to the atrophy of the paravertebral muscles [8, 9]. The aspect of paraspinal muscle mass loss in CBP across different age groups and the development of localized spinal sarcopenia in spinal OA has not been adequately studied, despite its critical importance in developing rehabilitation and prevention strategies for this patient cohort.

The aim of this study was to identify spinal sarcopenia in individuals with spinal OA by assessing spinal muscle mass and analyzing its relationship with degenerative spinal changes across various age groups using magnetic resonance imaging (MRI) of the lumbar spine.

## Patients and methods

The study was conducted at Pirogov Russian National Research Medical University (RNRMU) in collaboration with the Radiology Department of City Clinical Hospital No. 1 named after N.I. Pirogov, Moscow, during 2020–2021. The work was approved by the local ethics committee of RNRMU. All patients signed informed consent to participate in the study. The study included 90 patients with OA, divided into three age groups based on the WHO criteria (2012): young adults (Group I, n=35), middle-aged adults (Group II, n=30), and older adults (Group III, n=28) (Table 1).

**Table 1.** Demographic characteristics of the patients. Data shown as median and interquartile range.

|  | **Young adults**<br>**I group, n=35** | **Middle-aged adults**<br>**II group, n=30** | **Older adults**<br>**III group, n=28** |
| --- | --- | --- | --- |
| Age, years | 36.00 [30.00-42.00] | 50.00 [46.50-56.25] | 66.00 [62.50-71.00] |
| Males, n (%) | 18 (51.4%) | 11 (36.7%) | 18 (64.3%) |
| Females, n (%) | 17 (48.6%) | 19 (63.3%) | 10 (35.7%) |

All patients with previously diagnosed spinal OA suffered from CBP, so they were examined at the department. As part of the examination, all participants underwent lumbar spine MRI to identify clinically significant structural and anatomical changes, such as intervertebral disc herniation with neural compression, lumbar/foraminal stenosis, spondylolisthesis, and others. Physical examination included assessment of body mass index (BMI), pain intensity using a visual analog scale (VAS, 0–100 mm), classification of back pain (BP) as acute or chronic based on the International Association for the Study of Pain (IAPS, 2021) criteria, and the degree of functional limitation in the lumbar spine using the Backache Index (BAI). Neurological complications, including radicular syndrome and signs of neurological deficits (sensory and/or motor impairments, pelvic organ dysfunction), were also evaluated.

Inclusion criteria comprised patients with MRI-confirmed spinal OA, exhibiting 4th–5th stage intervertebral disc degeneration according to the Pfirrmann classification and presence of disc herniation. Exclusion criteria included specific causes of CBP, such as vertebral compression fracture, absolute lumbar canal stenosis, tumor or metastasis to the spine/spinal cord, infectious spondylodiscitis, and inflammatory rheumatologic spondyloarthritis.

MRI of the lumbar spine was performed using a 1.5 Tesla scanner (Toshiba, 1.5 T, Japan). Patients were positioned supine with a cushion under the head and knees, maintaining a neutral lumbar spine position. The imaging protocol included T1- and T2-weighted sequences in two planes: sagittal and axial. Axial images analyzed at a level parallel to the upper end plate of the third lumbar vertebra (L3). Analysis of spinal muscle mass was performed at the L3 level in the axial plane, calculating the total cross-sectional area (CSA) of paired spinal muscles (Sm, cm²), including the psoas major, erector spinae, and quadratus lumborum muscles, using manual contouring with the ‘Curved Contour’ tool [10]. Similarly, the CSA of the L3 vertebral body at the superior endplate (Sv, cm²) was measured (Figure 1).

**Figure 1.**
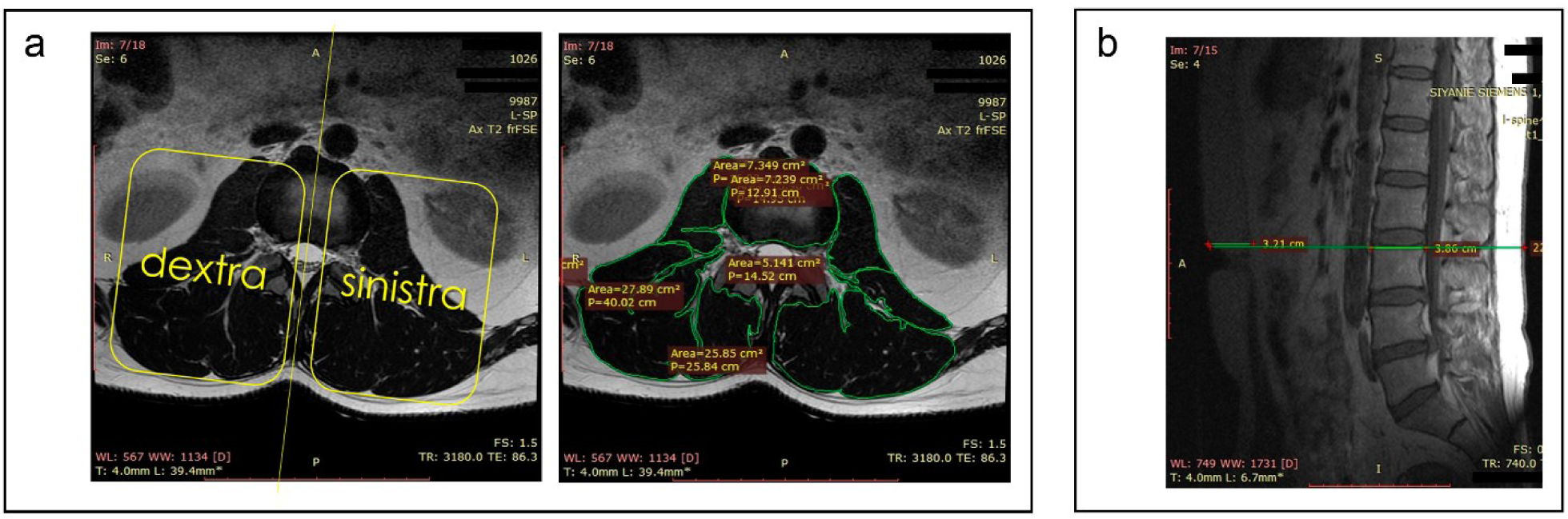
Measurement of spinal parameters (the thickness of the body of the third lumbar vertebra, torso thickness at the lumbar level) and the mass of spinal muscles. a. Assessment of the dimensions of the L3 vertebral body and trunk thickness at the lumbar level. b. Contouring of the three groups of paraspinal muscles: the psoas major, erector spinae, and quadratus lumborum, along with the L3 vertebral body. The sample/patient IDs are known only to the research group and not to anyone outside of it.

Considering the variability in absolute muscle mass values among patients with differing anthropometric characteristics and the potential for measurement error due to image scaling differences, muscle mass indices were normalized relative to vertebral body CSA. The muscle-vertebral index (MVI) was calculated using the formula: the sum of bilateral paraspinal muscle CSA divided by the CSA of the L3 vertebral body (MVI = [Sm_dextra + Sm_sinistra]/Sv). To detect asymmetric paraspinal muscle development in cases of scoliotic spinal deformities and assess its contribution to muscle-tonic syndrome, the ratio of left-to-right paraspinal muscle CSA was calculated (Figure 1a).

Other MRI parameters analyzed included intervertebral disc degeneration stage using the Pfirrmann classification (1–5 stages, 2001) [11] and sagittal trunk thickness at the L3 level in horizontal position (Figure 1b).

## Results

The average intensity of lower back pain was higher in young and elderly patients, compared to middle-aged individuals (p = 0.0528). The average degree of functional limitations in the lumbar spine across all examined groups corresponded to a high level (above 0.7 on the BAI) and showed no statistically significant differences between groups (p > 0.05) (Table 2). The BAI positively correlated with pain intensity on the VAS in all three groups (r = 0.552, p = 1.92 × 10^−6^) (Figure 2a).

**Figure 2.**
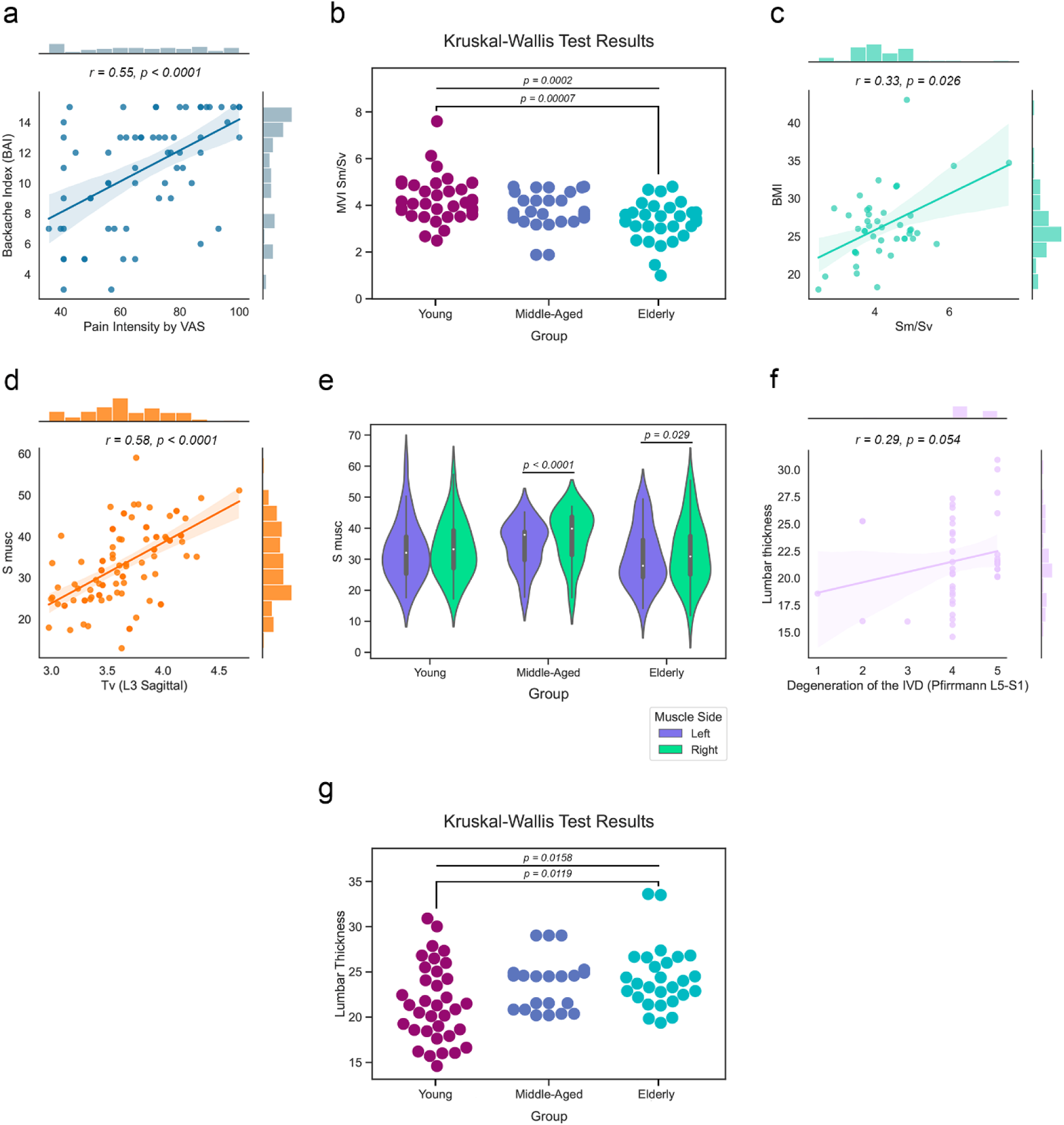
a. Positive correlation between BAI and pain intensity on the visual analog scale (VAS). b. Mean Muscle-Vertebral Index (MVI) values in patient groups. c. Positive correlation between BMI and MVI. d. Positive correlation between vertebral body thickness and spinal muscle area. e. Differences in the total area of the three spinal muscles are present only on the right side between age groups (Kruskal-Wallis test), with asymmetry observed in the middle-aged and elderly groups (Wilcoxon test). f. Trunk thickness at the lumbar level (L3 vertebra) in age groups. g. Positive correlation between the stage of intervertebral disc degeneration according to Pfirrmann at the L5-S1 level and lumbar thickness.

**Table 2.**
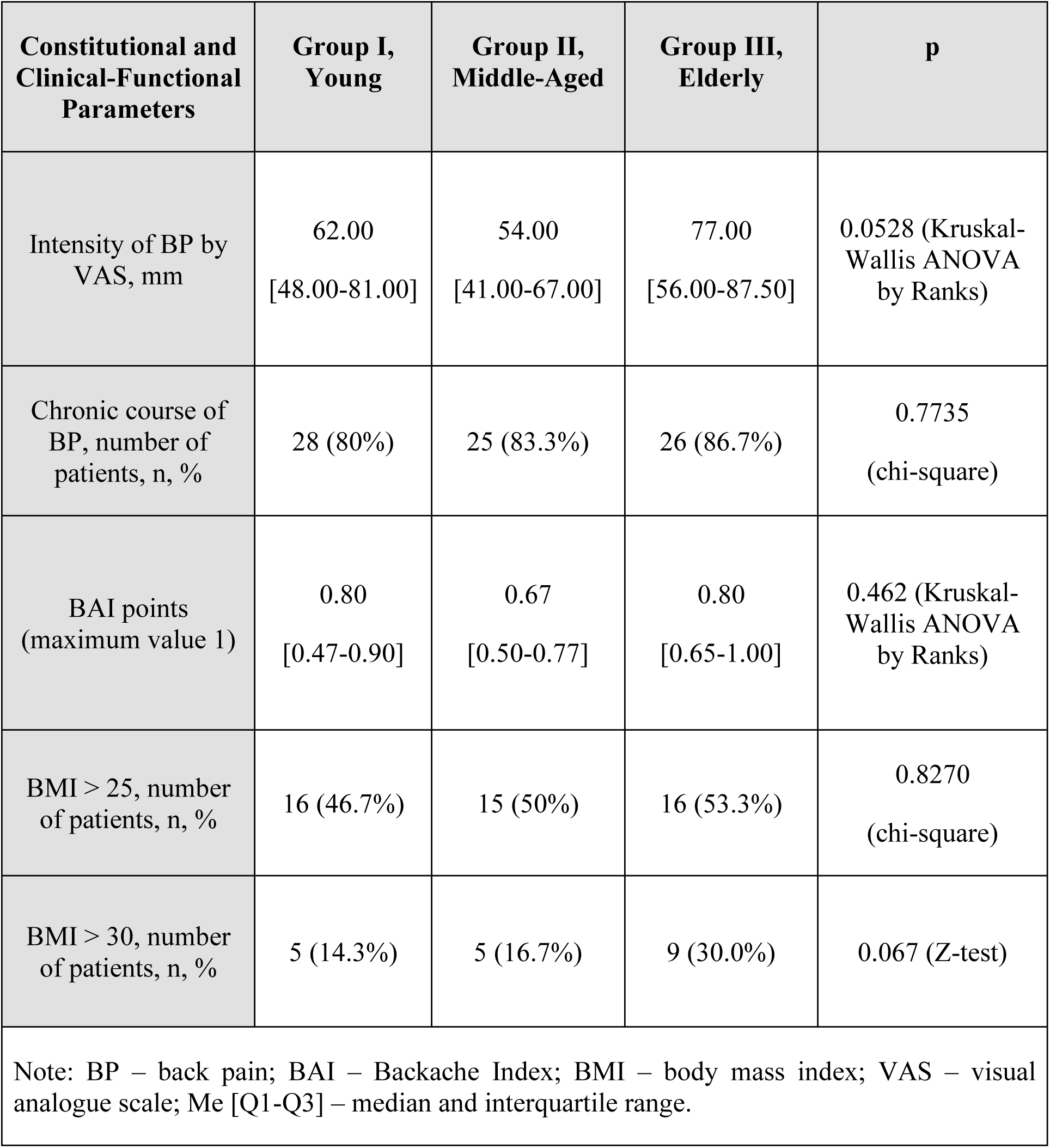
Constitutional and Clinical-Functional Data of Patients.

In the overwhelming majority of cases (>80%), BP was chronic. Half of the patients in each group were overweight, while 30% of elderly patients were obese, compared to young and middle-aged individuals (14.3% and 16.7%, respectively, p = 0.067).

The paraspinal muscle mass, as measured by the mean MVI values, was significantly lower in middle-aged (p = 0.025) and elderly individuals (p = 0.0001) compared to young individuals (Table 3, Figure 2b), indicating an age-related decline in muscle mass despite a statistically significant increase in the average cross-sectional area of the 3rd lumbar vertebra (Sv) was observed across the groups. This finding suggests the development of spondylosis and the growth of marginal osteophytes with age as manifestations of spinal OA.

**Table 3.** Muscle-Vertebral Index (MVI) and Cross-Sectional Areas of Muscles and the Body of the 3rd Lumbar Vertebra in the Study Groups.

| MVI and Cross-Sectional Areas of Paraspinal Muscles and the Body of the 3rd Lumbar Vertebra | Group I<br>(Young) | Group II<br>(Middle-Aged) | Group III<br>(Elderly) | p I and II | p I and III | p II and III |
| --- | --- | --- | --- | --- | --- | --- |
| MVI Sm/Sv | 4.12 [3.60-4.93] | 3.66 [3.43-4.29] | 3.37 [2.93-3.75] | 0.0253 | 0.0001 | 0.0286 |
| Sv, cm<sup>2</sup> | 15.01 [13.98-16.37] | 18.76 [17.52-21.97] | 18.51 [15.32-20.49] | $4.750 \times 10^{-8}$ | 0.0003 | 0.3633 |
| MVI m. quadratus lumborum | 0.53 [0.44-0.68] | 0.47 [0.38-0.60] | 0.34 [0.27-0.48] | 0.0642 | $3.704 \times 10^{-5}$ | 0.0022 |
| MVI m. psoas major | 0.97 [0.83-1.27] | 0.83 [0.77-0.93] | 0.80 [0.68-1.06] | 0.0262 | 0.0054 | 0.5332 |
| MVI m. erector spinae | 2.52 [2.20-3.07] | 2.39 [2.31-2.76] | 2.19 [1.99-2.47] | 0.0898 | 0.0042 | 0.1464 |
| Thickness of the lower back | 21.30 [18.44-25.04] | 24.51 [20.83-24.90] | 23.70 [22.19-26.60] | 0.0400 | 0.0078 | 0.5147 |
| Note: MVI – muscle-vertebral index; Sv – cross-sectional area of the 3rd lumbar vertebra at the level of the upper endplate; Me [Q1-Q3] – median and interquartile range. |  |  |  |  |  |  |

Moreover, a positive correlation was identified between BMI and MVI (r = 0.333, p = 0.026), indicating an adaptive increase in spinal muscle mass with weight gain (Figure 2c). Additionally, a positive correlation was observed between vertebral body thickness and spinal muscle area (r = 0.576, p = 1.055 × 10^−9^) (Figure 2d), suggesting that muscle mass increases alongside the axial skeleton components, specifically vertebral bodies.

To determine which muscles experience the greatest area reduction with age, MVI was calculated, and the absolute cross-sectional areas of each pair of spinal muscles were compared. A statistically significant reduction in mass was found for the quadratus lumborum muscle in elderly individuals compared to both middle-aged (p = 0.0022) and young individuals (p = 3.704 × 10^−5^), for m. psoas major in middle and elderly compared to young (p = 0.0262 and p = 0.0054), for m. erector spinae in the elderly compared to the young (p = 0.0042). (Table 3). For m. quadratus lumborum, asymmetry was observed across all age groups (Table 4), being more pronounced in middle-aged individuals (p = 2.762 × 10^−6^) and young individuals (p = 0.0009), and less pronounced in elderly individuals (p = 0.0037). When comparing the total cross-sectional area of the three paraspinal muscles, differences between age groups was significant for the right side (p = 0.0252) (Figure 2e). This likely reflects compensatory hypertrophy on the right side of the trunk in all subjects.

**Table 4.** Asymmetry of the m. quadratus lumborum, m. psoas major and m. erector spinae on the Left and Right in Age Groups, calculated by the cross-sectional area. Wilcoxon signed-rank test, p < 0.05 only.

| Groups | Left, Me [Q1-Q3] | Right, Me [Q1-Q3] | p |
| --- | --- | --- | --- |
| S of m. quadratus lumborum |  |  |  |
| Group I<br>(Young) | 3.73 [2.60-5.34] | 4.34 [3.47-5.71] | 0.0009 |
| Group II<br>(Middle-Aged) | 4.22 [2.95-4.72] | 5.45 [3.51-6.65] | $2.762 \times 10^{-6}$ |
| Group III<br>(Elderly) | 2.64 [2.20-4.20] | 2.95 [2.45-5.29] | 0.0037 |
| S of m. psoas major |  |  |  |
| Group 1 | 7.41 [5.56-10.12] | 7.81 [6.03-10.12] | 0.0356 |
| Group 2 | 7.90 [5.84-10.54] | 9.03 [7.49-9.89] | 0.0216 |
| S of m. erector spinae |  |  |  |
| Group 2 | 22.10 [19.87-26.70] | 23.23 [17.88-26.64] | 0.0061 |

The trunk thickness at the lumbar level in the sagittal plane based on MRI data (patients lying on their backs during the examination) was statistically greater in elderly individuals compared to young individuals (p = 0.0119) (Figure 2f). Considering the prevalence of obesity in the elderly population, this finding may indirectly suggest a larger abdominal volume in this age group. This could indicate an increase in visceral fat, as well as a reduction in the tone of the anterior abdominal wall muscles due to age-related muscle tone loss and weight gain.

Weak positive correlation was found between the Pfirrmann grading stage of disc degeneration and lumbar thickness (r = 0.292, p = 0.05416) (Figure 2g), which indirectly suggests an influence of abdominal volume on disc degeneration and requires further confirmation.

## Discussion

The interplay between primary sarcopenia, associated with aging, and localized spinal sarcopenia in spinal OA is an area of active research [12]. The exacerbation of CBP in in this context is evident due to destabilization of the motion segment, formation of intervertebral disc extrusions, spondylolisthesis, and worsening of facet joint syndrome. Unfortunately, the existing diagnostic criteria for primary sarcopenia have limitations when applied to spinal sarcopenia [13, 14]. These limitations stem from the lack of a standardized protocol for assessing paraspinal muscle mass using imaging modalities such as CT, MRI, dual-energy X-ray absorptiometry, or bioimpedance analysis. Furthermore, equipment to measure the isokinetic strength and function of spinal flexor and extensor muscles separately is often unavailable, and their activation may induce BP.

Routine MRI can serve as a tool to assess paraspinal muscle mass loss through a simple and accessible measurement of the MVI using a single axial slice at the level of the third lumbar vertebra (L3), where all three paired paraspinal muscle groups are represented [15]. Unlike previous indices, the MVI does not require patient contact or individualized body parameter calculations such as BMI, height, or body surface area [10]. It can be readily evaluated by a radiologist. Our findings on the correlation between MVI and BMI, reflecting the adaptive increase in paraspinal muscle mass with body weight and axial spinal loading, further support the utility of this parameter.

It is well-established that the sources of nociceptive input in BP can include both osseous-cartilaginous components of the spinal motion segment, segmental spinal instability, and the musculoskeletal apparatus [16]. The significant contribution of myofascial syndrome to CBP underscores the need for studies addressing the tone, mass, and adaptive mechanisms of paraspinal muscles across age groups and in young office workers [17]. Previous studies have shown that trunk and abdominal muscles are functional components of the motion segments, stabilizing them during acute pain episodes and preventing vertebral displacement or disc slippage [18]. CBP, on the other hand, limits local lumbar spine mobility and reduces paraspinal muscle activity. A study by Lao et al., using kinetic MRI in 162 non-operated BP patients, revealed significant intervertebral movement restriction, progressing to ankylosis of the motion segment in Pfirrmann grade 5 intervertebral disc degeneration (IDD) [9].

Our findings, along with other studies, demonstrate age-related paraspinal muscle mass loss, with MVI values being significantly lower in older adults compared to younger and middle-aged individuals [10, 15, 19]. This phenomenon is likely influenced by generalized skeletal muscle atrophy in older adults. Interestingly, the loss of muscle mass is associated with an increase in vertebral body cross-sectional area, particularly at the support endplates, despite a direct correlation between MVI and the L3 vertebral cross-sectional area, as shown in our work. Large-scale studies on OA have long recognized aging as the strongest risk factor for bone degeneration, particularly in the spine [20].

Lumbar spondylosis, historically termed ‘deforming spondylitis’ or ‘intervertebral osteochondrosis’, is characterized by pathological endplate osteophyte formation, narrowing of intervertebral spaces, disc degeneration, and reactive changes in adjacent vertebral bodies [21]. Numerous studies have demonstrated a strong correlation between the expansion of vertebral body endplate surfaces and aging [22, 23]. According to some authors, osteophyte formation represents an adaptive mechanism in response to spinal instability and reflects remodeling of the motion segment in response to altered spinal biomechanics [24]. However, these bony projections, encroaching on the spinal canal or intervertebral foramina, may restrict motion, immobilize the motion segment, and exacerbate axial spine pain by penetrating adjacent organs, tissues, or ligaments [25].

In our work, the asymmetry of all three groups of paired paraspinal muscles with predominance of the right side was revealed, consistent with findings in similar studies of patients with spinal OA [26]. Interestingly, asymmetry was also noted in spinal muscle strength endurance in 7–8-year-old girls with asymmetric posture and grade 1 scoliosis, with reduced strength on the left side [27]. Office workers are affected by myofascial pain syndrome, including the quadrangular lumbar muscle involvement [28]. The quadratus lumborum muscle originates at the posterior iliac crest and iliolumbar ligament and inserts into the medial border of the 12th rib and the transverse processes of the L1–L5 vertebrae. It is innervated by muscular branches of the lumbar plexus (Th12, L1–L5) [29]. Its fibers are frequently involved in myofascial pain syndrome, with active and latent trigger points.

This deep lumbar muscle, located beneath the erector spinae muscle, can be accessed manually in the lateral recumbent position for trigger point palpation in cases of myofascial pain syndrome [30]. The quadratus lumborum serves critical functions, including (1) trunk stabilization in an upright position during bilateral contraction (a global stabilizer); (2) lowering the 12th ribs during deep abdominal exhalation; and (3) lateral trunk flexion during unilateral contraction. Recognizing its pivotal role in paraspinal muscles mass loss, rehabilitation programs for patients with chronic lumbar pain and spinal OA should include specific exercises targeting the dynamic and isometric engagement of these muscles bilaterally, along with additional left-side-specific exercises. Such exercises include lateral trunk flexion (standing side bends, or lateral lifts of the head and torso in the lateral recumbent position) and respiratory exercises involving deep exhalation and breath-hold repetitions for isometric engagement of both quadratus lumborum muscles [31].

The greater trunk thickness observed in older adults compared to younger and middle-aged individuals in our study likely reflects abdominal volume increase with age. This parameter was assessed in a supine position, minimizing gravitational effects. Previous studies have identified correlations between abdominal volume, lumbar lordosis angle, and IDD severity [32]. For patients with lower BP and spinal OA, therapeutic exercise regimens should include abdominal muscle activation, along with weight reduction efforts. In younger patients, these exercises can include direct and lateral trunk flexion and pelvic tilts. For older patients with reduced functional status, breathing exercises and abdominal muscle tone activation throughout the day – such as alternating abdominal wall contractions while sitting, standing, or walking – can help develop new motor patterns.

Notably, maximum efficiency in individual muscle function is achieved through targeted exercise regimens activating multiple core stabilizers. These static and statodynamic exercises improve proprioception and coordination, particularly important for older adults, and form the foundation for strengthening the trunk muscle corset in comorbid patients and preventing CBP in younger individuals [31]. Furthermore, non-surgical approaches focusing on lumbar spine stabilization emphasize the activation of spinal muscles (including the m. quadratus lumborum) and abdominal muscles, providing relief for intervertebral discs, facet joints, and ligaments [33]. Comprehensive rehabilitation programs, incorporating manual techniques, dynamic and isometric exercises, and breathing practices, are critical for enhancing spinal muscle function and posture normalization, including among office workers, to reduce the incidence of new pain episodes and BP onset in all age groups [34].

## Conclusions

The mass of spinal muscles in patients with spinal OA decreases with age, which is reflected in the reduction of muscle-to-vertebra ratio values on MRI. The obtained data indicate the formation of spinal sarcopenia and its probable contribution to the development of OA with aging. The asymmetry of all three groups of paired paraspinal muscles was revealed (with greater mass on the right side than on the left). Lumbar thickness, as well as body mass, increased with age in patients with spinal OA, which may indicate reduced tone in the abdominal muscles within the context of age-related sarcopenia, exacerbating the clinical course of the disease. The obtained data allow for the formulation of priority tasks for maintaining the spinal framework across all age groups to prevent spinal degeneration and CBP. The non-surgical approach, including activation of spinal and abdominal muscles, as well as comprehensive rehabilitation programs aimed at strengthening muscles and normalizing posture, plays a key role in preventing new pain episodes.

## Study Limitations

This study did not evaluate spinal muscle function due to patient assessments being conducted during exacerbation phases with significant BP. Currently, there is no established ‘gold standard’ for assessing spinal sarcopenia. Comprehensive evaluation of spinal sarcopenia using criteria from the European and Asian societies for sarcopenia research was not feasible, given the absence of assessments for muscle strength and function. Furthermore, the study did not include an evaluation of skeletal musculature relevant to spinal function, such as the abdominal muscles and pelvic floor muscles, commonly referred to as ‘core strength’. Such an approach may represent a valid and promising direction for assessing the spinal muscular framework in future studies.

## Authors’ Contributions

All authors made significant contributions to the study’s preparation, reviewed, and approved the final manuscript version before publication.

N. G. Pravdiuk – literature review, study design, data analysis, and interpretation; A.V. Novikova – literature review, clinical data collection, data analysis and interpretation, and preparation of graphical materials; N. A. Shostak – approval of study design, research consultation, and manuscript editing; A.A. Klimenko – research consultation and manuscript editing; E.S. Pershina – provision of MRI images, expert assessment of imaging and statistical data; A.A. Muradyants – research consultation and manuscript editing; A.A. Buianova – preparation of figures and manuscript editing.

## Data Availability

All data produced in the present study are available upon reasonable request to the authors.

## Acknowledgements

The authors thank Y.S. Zhulina, clinical resident at I.M. Sechenov First Moscow State Medical University, for her contributions to MRI image analysis, figure preparation, and MVI calculation.

## Ethics

The study design was approved by the local ethics committee (LEC) of Pirogov Russian National Research Medical University (protocol excerpt No. 208 of the LEC session dated May 17, 2021).

